# Assessing Depression Severity Through a Passive Digital Mood Marker

**DOI:** 10.64898/2025.12.20.25341076

**Authors:** Antony Perzo, Nour Lachtar, Fabrice Boulet, Renaud Séguier, Clara Roux-Pertus

## Abstract

**Objective:** Effective management of Major Depressive Disorder (MDD) is limited by reliance on episodic, subjective clinical scales. Passive digital phenotyping offers a potential solution for continuous, objective monitoring. We aimed to assess the concurrent validity of a novel digital biomarker—the Facial Affect Dynamics–derived Depression Severity (FADS) score—against the Patient Health Questionnaire-9 (PHQ-9).

**Methods:** We conducted an interim analysis of the EMC2FR study (NCT06860165), a prospective, multicenter cohort study in France. Seventeen outpatients with unipolar MDD used the EmoDTx smartphone application, which passively analyzes facial expressions via the front-facing camera during routine device use. We analyzed data from Week 2 (W2) and Week 6 (W6) visits. The association between the 14-day passive FADS score and the PHQ-9 was estimated using Spearman rank correlation with participant-level bootstrapping to account for repeated measures.

**Results:** The analysis included 34 person-visit observations from 17 participants (70.6% women; mean age 42.7 years). We observed a strong, positive concurrent correlation between FADS and PHQ-9 scores (ρ = 0.65; 95% CI, 0.50–0.87; P =.01). Furthermore, within-participant changes in FADS scores from W2 to W6 were strongly correlated with changes in PHQ-9 scores (ρ = 0.67, P =.02).

**Conclusions:** Preliminary results suggest that passive monitoring of facial affect dynamics is associated with clinically relevant variation in depression severity. These findings support the ongoing EMC2FR study to validate this marker as a tool for longitudinal depression monitoring.

## Introduction

Effective management of Major Depressive Disorder (MDD) is challenged by reliance on subjective, episodic clinical scales, which may not fully capture day-to-day symptom fluctuations. Digital phenotyping, leveraging passively collected signals from personal devices, may complement questionnaires by providing objective, continuous biomarkers that could enhance depression monitoring.^1^ Prior work has shown associations between depressive symptoms and passively collected data, such as GPS-based mobility patterns and keystroke dynamics.^2,3^ While passive facial analysis has shown moderate accuracy for diagnostic classification,^4^ its validation for longitudinal symptom monitoring remains a critical gap. We report interim findings from an ongoing, prospective, multicenter study (EMC2FR; NCT06860165) on the concurrent association between a novel facial affect dynamics–derived depression severity score (FADS) and the Patient Health Questionnaire-9 (PHQ-9).^5^

## Methods

### Study Design and Participants

This analysis is from a prospective cohort study conducted at three sites in France, approved by CPP Sud-Est I; all participants provided written informed consent. We included 17 outpatients (mean (SD) age, 42.7 (12.7) years; 12 (70.6%) women; mean (SD) baseline PHQ-9 score, 11.8 (4.9)) diagnosed with unipolar MDD. Per protocol, the only time points with both the passive FADS and the PHQ-9 were the Week 2 (W2) and Week 6 (W6) remote visits, yielding 34 person-visits.

### Digital Phenotyping (EmoDTx)

The EmoDTx application (Emobot) passively analyzes facial expressions from the front-facing camera during device use with an in-house deep learning model that quantifies patterns and dynamics of emotional expressivity, generating a continuous marker of depression severity. For each visit, the FADS summarizes data from the 14 calendar days immediately preceding the PHQ-9 administration (aligned with its two-week recall period), with higher values indicating greater depression severity.

### Statistical Analysis

Primary analysis estimated the monotonic association between FADS and PHQ-9 at the visit level using the Spearman rank correlation. Because observations are clustered within participants (2 per person), all interval estimates used participant-level bootstrapping: we resampled participants (G=17) with replacement, retained both W2 and W6 rows for sampled participants, and recomputed ρ for each bootstrap sample (B = 10,000). We report bias-corrected and accelerated (BCa) 95% CIs. For hypothesis testing of *H0* : *ρ = 0*, we used a cluster-wise permutation test (B = 50,000 shuffles): we randomly permuted the mapping of PHQ-9 participant blocks to FADS participant blocks (preserving each participant’s W2/W6 pair) and recomputed ρ to form the null distribution. This preserves within-participant dependence. Secondarily, to assess the marker’s sensitivity to change, we calculated the within-participant change scores for both measures from W2 to W6 (ΔFADS and ΔPHQ-9) and computed the Spearman correlation between these change scores (N=17).

## Results

We observed a strong, positive concurrent correlation between the FADS and PHQ-9 scores across the 34 person-visit observations (ρ = 0.65; 95% CI, 0.50–0.87; *P* =.01) (Figure 1). This indicates that higher FADS values were associated with higher self-reported depressive symptom severity. Furthermore, there was a strong positive correlation between the within-participant change in FADS and the change in PHQ-9 from W2 to W6 (ρ = 0.67, *P* =.02).

**Figure 1.**
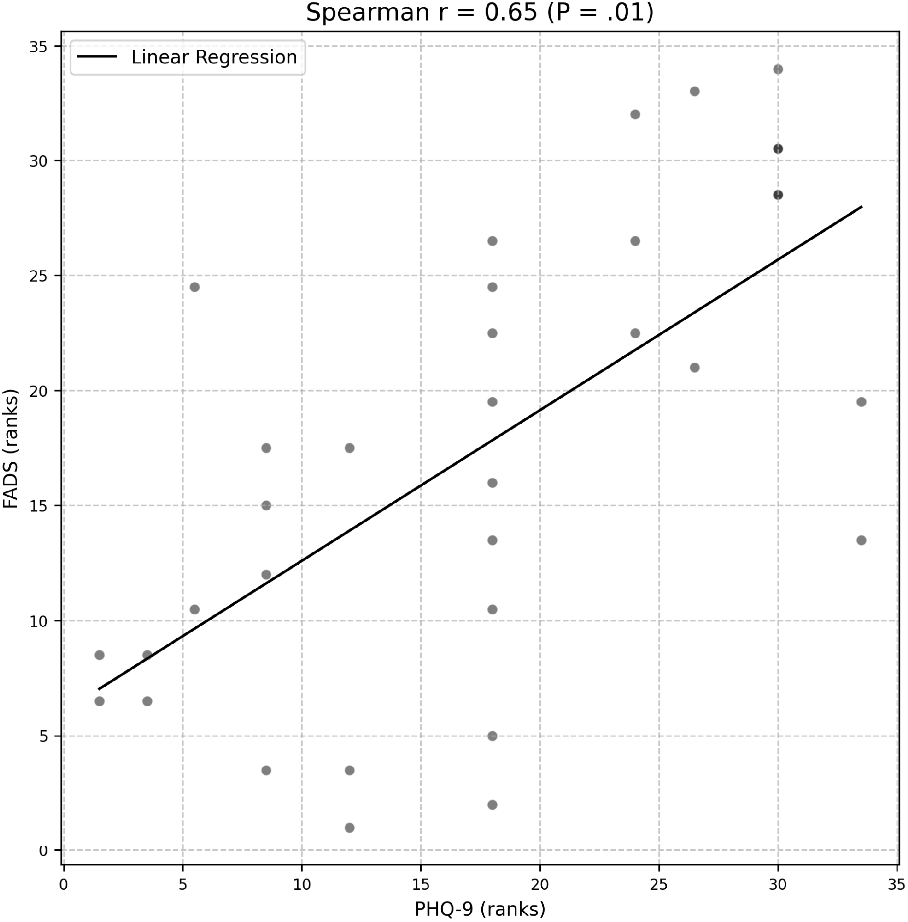
Rank–rank visualization of the concurrent association between FADS and PHQ-9 at W2 and W6 (n=34 person-visit observations from 17 participants). Points show mid-ranks (average method) for each measure. The line shows a least-squares fit to the ranks. Spearman ρ = 0.65 (BCa 95% CI, 0.50–0.87); P =.01 (cluster-wise permutation).

## Discussion

These preliminary findings suggest that passive monitoring of facial affect dynamics is associated with clinically relevant variation in depression severity. Primary limitations include the interim nature of the analysis and the small sample (17 participants; 34 person-visits, 17% of the planned N=98), which limits statistical power and precision. In addition, the limited number of time points per participant restricts our ability to model high-frequency temporal dynamics. Furthermore, these results are compared with a self-report measure; validation against planned gold-standard, clinician-rated endpoints is essential.

## Conclusion

These encouraging early results support continued enrollment and completion of the EMC2FR study to establish the validity of this depression severity marker.

## Data Availability

Upon completion of the full trial, deidentified participant data will be made available upon reasonable request to the corresponding author.

## ARTICLE INFORMATION

### Author Contributions (CRediT)

- Antony Perzo: Conceptualization; Methodology; Formal analysis; Investigation; Writing – original draft; Writing – review & editing; Supervision; Project administration
- Nour Lachtar: Conceptualization; Formal analysis; Data curation; Writing – review & editing; Visualization
- Fabrice Boulet: Methodology; Investigation; Writing – review & editing; Supervision
- Renaud Séguier: Writing – review & editing
- Clara Roux-Pertus: Methodology; Investigation; Writing – review & editing; Supervision; Project administration

### Additional Contributions

We thank the participants of the EMC2FR study.

